# Improving Responsiveness in Game-based Cognitive Assessment for Mild Cognitive Impairment

**DOI:** 10.1101/2025.07.25.25332206

**Authors:** Juhyeon Lee, Aurora James-Palmer, Isaac Heitmann, Allison Bierly, Jean-Francois Daneault, Sunghoon Ivan Lee

## Abstract

Mild Cognitive Impairment (MCI) affects up to 20% of older adults and often progresses to dementia. While brief cognitive screening tools like the Montreal Cognitive Assessment (MoCA) can aid in early detection and monitoring, their reliance on trained clinicians and susceptibility to test anxiety limit accessibility and ecological validity. Game-based cognitive monitoring presents a promising alternative, yet its sensitivity to cognitive changes in individuals with MCI remains underexplored. This study introduces an analytic pipeline for screening cognitive decline using Neuro-World, a serious gaming platform featuring six adaptive subgames that assess cognitive abilities through metrics such as accuracy and response time. Over 12 weeks, ten participants with MCI completed 24 game sessions. Gameplay data were analyzed using correlation-based feature selection and machine learning models to estimate cognitive function and track longitudinal changes. Results showed strong correlations between game-based assessments and clinician-administered MoCA scores (*r* = 0.71), as well as with longitudinal cognitive changes (*r* = 0.80). These findings highlight the potential of game-based cognitive assessments to provide self-administered, ecologically valid screening for cognitive decline in MCI, supporting early detection in aging populations.

## I. INTRODUCTION

Mild Cognitive Impairment (MCI) is characterized by a level of cognitive decline that lies between the expected changes associated with normal aging and the early stages of dementia [1]. Individuals with MCI often experience subtle cognitive impairments, such as increased forgetfulness or a reduced attention span, while their ability to perform activities of daily living remains intact or only minimally affected [1]. The prevalence of MCI among individuals aged 60 years and older is estimated to range from 15 to 20%, reflecting its frequent occurrence and increasing prevalence as the aging population grows [2].

MCI progresses to dementia at an annual rate of 8–15%, and individuals with MCI face a five-to ten-fold increased risk compared to cognitively healthy individuals [2], [3]. Given the profound impact of dementia on quality of life and independence, frequent and reliable screening of cognitive decline is crucial for monitoring progression in individuals with MCI [1].

Current clinical screening for cognitive impairment primarily relies on standardized tools such as the Mini-Mental State Examination (MMSE) [4] and the Montreal Cognitive Assessment (MoCA) [5]. While these tests are more practical and time-efficient than comprehensive neuropsychological batteries—the gold standard for cognitive evaluation [5], they still require administration by trained clinicians, which limits accessibility, as assessments must be conducted through inperson visits or, at minimum, remote video or phone consultations [6], [7]. Additionally, the structured clinical setting may induce anxiety, potentially affecting performance and reducing the ecological validity of the assessments [8].

Recent advancements in mobile health technologies have introduced serious games as a promising tool for self-administered, ecologically valid cognitive assessments [9]. Many studies have demonstrated that features extracted from gameplay performance data correlate with standardized cognitive test scores [9]–[11]. However, few prior studies have explored the responsiveness of game-based cognitive assessments in capturing longitudinal changes in cognitive function.

In this paper, we propose an analytic pipeline for game-based assessment of cognitive impairments and disease progression in individuals with MCI. We utilized a serious game called Neuro-World, which was used by ten participants with MCI over a 12-week period. Building on our prior study demonstrating that game data features from Neuro-World effectively estimate cognitive impairments in chronic stroke patients [10], we hypothesized that these features could accurately assess cognitive impairments and track changes in cognitive function over time in individuals with MCI. We employed a correlation-based feature selection method to identify features that capture both cross-sectional and longitudinal aspects of cognitive impairments. Using the selected features, we trained machine learning models to correlate with clinician-evaluated cognitive scores, thereby demonstrating the convergent validity and responsiveness of the proposed approach. These preliminary findings suggest that serious game-based cognitive assessments show potential as a practical tool for screening and monitoring cognitive decline in individuals with MCI and for supporting early detection in aging populations.

## II. METHODS

### A. Overview of Neuro-World

Neuro-World (Woorisoft Ltd., South Korea) is a serious game designed to assess cognitive impairment levels [10]. It consists of six subgames: Finding Directions, Remembering Animals, Remembering Sequences, Counting Animals, Finding Animals, and Matching Shapes, as illustrated in Fig. 1. Detailed descriptions of each subgame have been provided in our prior study [10]. While the previous study utilized the Korean version of Neuro-World, the present study employed its English version to accommodate participants in the United States. The difficulty of each subgame starts at level 1 and gradually increases up to level 20, the most challenging level. Game levels increase by one after three consecutive correct answers and decrease by one after two consecutive incorrect answers, ensuring adaptive difficulty based on user performance.

**Fig. 1.**
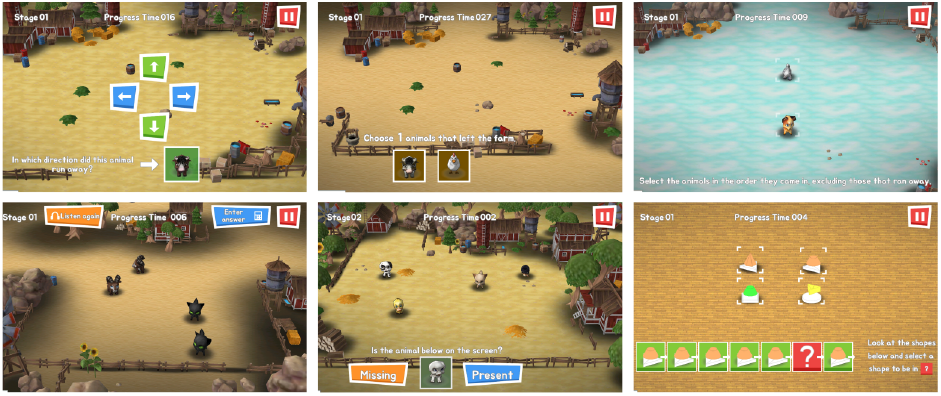
Screenshots of six subgames in Neuro-World. Top row: Finding Directions, Remembering Animals, and Remembering Sequences. Bottom row: Counting Animals, Finding Animals, and Matching Shapes.

Users played Neuro-World on a tablet PC using touch-based input. Gameplay data were wirelessly transmitted to a server and included detailed performance metrics for each question item, such as game level, answer correctness, time taken to answer each item, time taken to make the first touch, number of missed touches, and number of missed touches near the target answer. These performance metrics were designed to capture accuracy, reaction time, and patterns of interaction that could reflect patients’ cognitive abilities.

### B. Data Collection

Twelve participants with MCI were recruited across the United States for this remotely conducted study. Given that individuals with MCI are often undiagnosed, eligibility was determined using the MoCA. Participants scoring between 17 and 25 were included, based on prior research defining this range as indicative of MCI [12]. To ensure participants could engage with the tablet-based game and communicate over videoconferencing, inclusion criteria required: 1) minimal technological literacy, defined as the ability to operate a tablet independently, and 2) access to a computer with video and audio communication capabilities. After recruiting the first participant, an additional criterion was introduced, restricting eligibility to individuals aged 55 years or older. This adjustment aligned with the study’s focus on older adults, as MCI is predominantly observed in this population. As a result, the mean and standard deviation of participants’ ages were 63.9 ± 10.6 years (range: 38-78). Exclusion criteria were established to rule out other potential causes of cognitive impairment besides MCI, such as traumatic brain injury or a formal diagnosis of dementia. Participants were also required to have no significant upper-limb motor impairments, uncorrected vision or hearing issues, or involvement in therapist-supervised cognitive training programs, ensuring they could fully engage with the game. The study was approved by the Institutional Review Board at the University of Massachusetts Amherst (#2585), and all participants provided written informed consent.

Participants were instructed to play the Neuro-World game twice a week for 12 weeks, completing a total of 24 sessions. Each session lasted 30 minutes, with each subgame lasting five minutes. Participants received a tablet PC (M10, Lenovo) preloaded with Neuro-World, shipped to their homes. Before starting the gameplay, a tutorial session was conducted via videoconferencing (i.e., Enterprise Zoom) to train participants on using the tablet PC and playing the game. For further data analysis, two participants who completed less than 50% of the total sessions were excluded due to insufficient game data for estimating cognitive function. After this exclusion, the remaining participants completed an average of 22.5 ± 2.16 sessions.

To validate the game-based cognitive assessments against standard cognitive test scores, trained researchers administered standardized assessments via Zoom, including videoconferencing versions of the MMSE [7] and MoCA [6], both before and after the 12-week gameplay period. Only MoCA scores were used for analysis, as the MMSE exhibited low sensitivity in detecting MCI and showed a ceiling effect [5]. The MoCA is scored on a 0–30 scale, with higher scores indicating better cognitive function. The mean baseline MoCA score was 24.7 ± 2.69 (range: 18–28), while the mean post-game score was 22.4 ± 3.10 (range: 17–28).

Baseline assessments were intended to occur within a week before the start of gameplay, and post-game assessments within a week after the final session, ensuring that gameplay data aligned with MoCA-based cognitive function. However, logistical challenges, including scheduling conflicts and tablet shipping delays, introduced some variability. The mean interval between the baseline assessment and the start of gameplay was 8.3 ± 5.4 days (range: 0–18 days), while the gap between the end of gameplay and the post-game assessment averaged 7.7 ± 4.3 days (range: 1–17 days).

### C. Game Feature Extraction

We hypothesized that gameplay data within a specific time-frame can be used to screen cognitive impairments. Based on the assumption, baseline cognitive scores were estimated using game data collected after the baseline assessment, while post-game cognitive scores were estimated using data collected before the post-game assessment, as illustrated in Fig. 2.

**Fig. 2.**
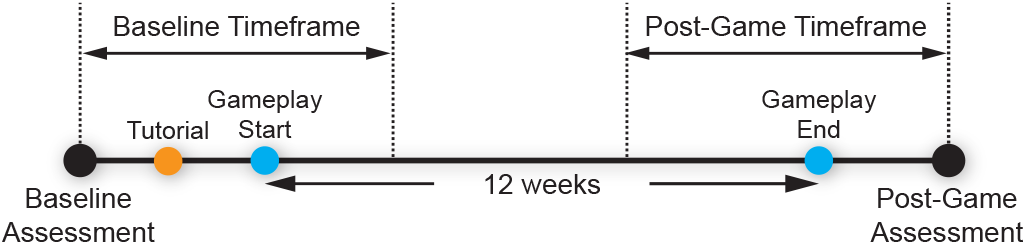
Data collection timeline and data inclusion timeframes for estimating baseline and post-game cognitive scores.

To determine the optimal timeframe for including game data in estimating cognitive impairments, a data-driven approach was employed. This approach accounted for potential cognitive changes over time in individuals with MCI as well as their responses to the game. The method aimed to identify the timeframe that maximized correlations between the extracted features and MoCA scores. The search range was set between 18 and 42 days, ensuring at least two gameplay sessions per participant and covering half of the 12-week gameplay period. Based on this method, 26 days of gameplay data were used to estimate baseline cognitive scores, and 24 days were used to estimate post-game cognitive scores. However, due to varying time gaps between the assessments and the start or end of gameplay, the number of sessions within these timeframes varied slightly across participants.

Game data within the determined timeframes were used to extract features designed to capture cognitive impairments. These features included game level, answer correctness, answer duration, first touch duration, missed touch counts, and target missed touch counts. For each session, features from all question items, except answer correctness, were aggregated using the mean and standard deviation. Answer correctness was aggregated into answer accuracy, defined as the percentage of correct answers within each session. Furthermore, features for each subgame within a session were aggregated using the mean, standard deviation, and answer accuracy. However, two subgames—i.e., Finding Directions and Remembering Sequences—were excluded from subgame-specific feature extraction due to insufficient data caused by low compliance among some participants and technical errors during gameplay. Additionally, missed touch counts and target missed touch counts were not extracted for subgame-specific feature extraction because several subgames did not record any missed touches, resulting in insufficient data for aggregation. Across all sessions, features were further aggregated using a range of statistical measures, including the mean, median, standard deviation, interquartile range, range, minimum, maximum, 10th percentile, and 90th percentile, resulting in a total of 351 derived features.

### D. Feature Selection and Machine Learning Models

Feature selection was performed using Correlation-based Feature Selection (CFS) to identify features with the strongest correlations to clinician-administered cognitive scores while minimizing redundancy among the selected features [13]. Unlike the conventional CFS approach, which considers only cross-sectional correlations between target cognitive scores and features, we extended the method by incorporating both cross-sectional and longitudinal correlations. The extended algorithm identified a subset of features by maximizing its merit, calculated using Equation (1):

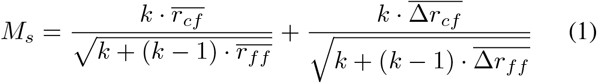

where *M*_*S*_ represents the merit of a feature subset *S* containing *k* features, 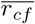 is the average correlation between features in subset *S* and the target scores, and 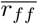 is the average intercorrelation between features within subset 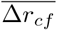 is the average correlation between changes in features within subset *S* and changes in the target scores, and 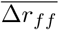 is the average intercorrelation of changes between features in subset *S*.

To prevent overfitting, the maximum number of features in the subset was restricted to 10, ensuring the number of selected features remained smaller than the number of participants. Spearman correlation was used for the merit calculations.

The selected features were then used to train six machine learning models: Lasso regression, linear regression, Gaussian Process regression, Support Vector Machine, Random Forest, and Adaboost. These models were trained to estimate baseline and post-game MoCA scores using the extracted features. To evaluate model performance, a leave-one-subject-out crossvalidation (LOSOCV) approach was employed. This method ensured that each participant’s data was treated as the test set once, while data from the remaining participants formed the training set. Hyperparameters for Lasso regression and support vector regression were optimized using a nested LOSOCV approach within the training data. For Lasso regression, the regularization parameter was selected from an exponentially spaced grid ranging from 10^−3^ to 10^3^ with 30 samples. For support vector regression, the *γ* and *C* hyperparameters were selected from an exponentially spaced grid ranging from 10^−3^ to 10^3^ with 7 samples each.

## III. RESULTS and Discussions

Among the 351 extracted features, an average of eight features per iteration were selected using a modified CFS approach. The best-performing machine learning model was Lasso regression. Lasso also reduced feature redundancy, utilizing an average of six features per iteration for regression.

Fig. 3 illustrates the cross-sectional association (*N* = 20) between game-based assessments and clinician-evaluated MoCA scores, combining both baseline and post-game data. The correlation was statistically significant, with Pearson’s *r* = 0.71, *p* < 0.001. Fig. 4 presents the longitudinal association (*N* = 10) between changes in clinician-evaluated MoCA scores and changes in game-based MoCA estimates. A strong correlation was observed, with Pearson’s *r* = 0.80, *p* = 0.006. These findings support the convergent validity and responsiveness of Neuro-World’s game-based cognitive assessment, demonstrating their ability to capture both cross-sectional cognitive function and longitudinal cognitive changes.

**Fig. 3.**
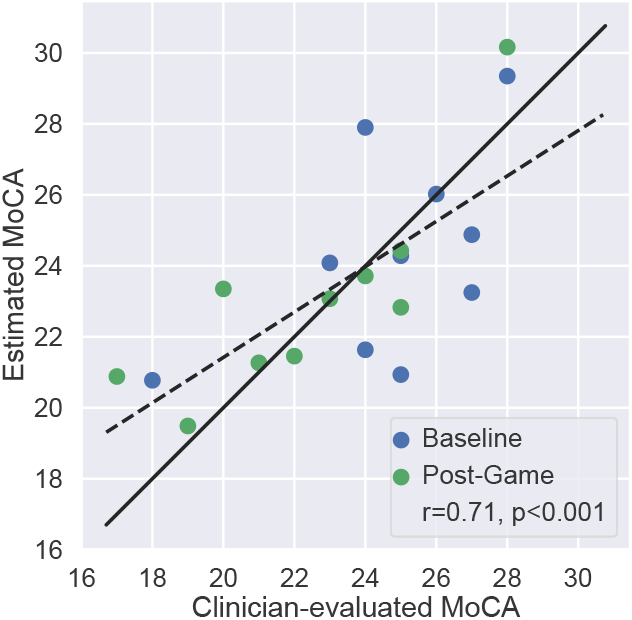
Clinician-evaluated MoCA vs. estimated MoCA by the proposed model. Baseline scores and post-game scores are represented by blue and green dots, respectively. The black solid line represents perfect agreements (*y* = *x*), while the black dotted line depicts the regression line for the data points.

**Fig. 4.**
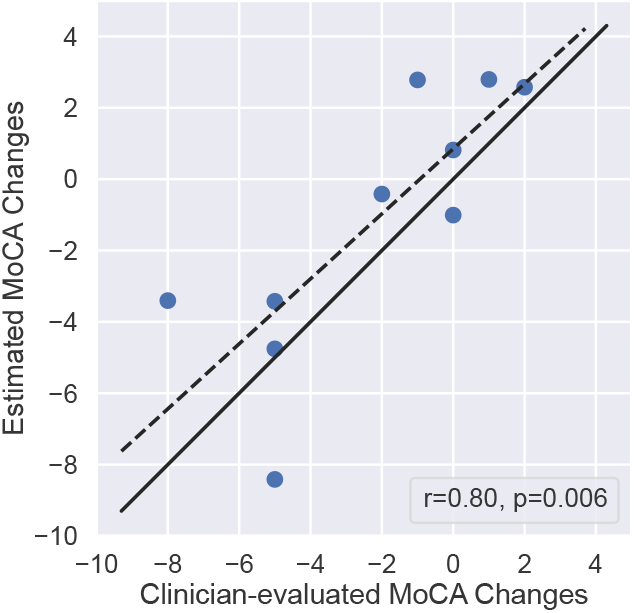
Clinician-evaluated MoCA Changes over time vs. estimated MoCA Changes. The black solid line represents perfect agreements (*y* = *x*), while the black dotted line depicts the regression line for the data points.

Among the frequently selected features, the variability of mean first touch duration across sessions showed the strongest correlations with MoCA scores (cross-sectional Pearson’s *r* = 0.72, *p* < 0.001) and longitudinal changes in MoCA scores (*r* = 0.69, *p* = 0.026). These positive correlations indicate that higher-functioning participants demonstrate greater flexibility in reaction times, suggesting an adaptive response to varying game difficulties. For instance, they responded more quickly to easier items while taking more deliberate actions on harder ones. This aligns with prior research showing that individuals with low-functioning MCI demonstrate reduced visuomotor adaptability and cognitive flexibility [14], [15].

There are limitations in this study. First, the small sample size limits the generalizability of the results, necessitating larger-scale studies to confirm robustness and applicability. Second, inconsistent adherence to gameplay schedules among participants affected the completeness of data collection. Future studies should explore strategies such as enhanced gamification or automated reminders to maintain participant engagement and support reliable data collection. Finally, while MoCA served as the primary reference measure, its limitations as an imperfect and coarse-grained ground truth call for the inclusion of more comprehensive cognitive assessments in future validations.

## IV. CONCLUSION

In conclusion, these preliminary findings showed the promise of serious games for screening and monitoring cognitive impairments in MCI. We envision that the game-based assessment enables frequent and fine-grained monitoring of cognitive changes without burdening clinicians and MCI patients with repeated clinic visits or videoconferencing sessions. Furthermore, the proposed method has the potential to be used as an accessible and practical tool for early detection and self-monitoring of cognitive decline in aging populations.

## Data Availability

All data produced in the present study are available upon reasonable request to the authors

## Notes

Research reported in this publication was supported by NIH/NIA Grant No. 1R21AG071988. The content is solely the responsibility of the authors and does not necessarily represent the official views of NIH.

### Competing Interest Statement

The authors have declared no competing interest.

### Clinical Trial

NCT04920123

### Funding Statement

This study was funded by NIH/NIA Grant No.
1R21AG071988

### Author Declarations

Institutional Review Board of the University of Massachusetts Amherst (#2585) gave ethical approval for this work

